# Using Hierarchical Clustering to Explore Patterns of Deprivation Among English Local Authorities

**DOI:** 10.1101/19006957

**Authors:** Steven L. Senior

## Abstract

**Background:** The English Indices of Multiple Deprivation (IMD) is widely used as a measure of deprivation of geographic areas in analyses of health inequalities between places. However, similarly ranked areas can differ substantially in the underlying domains and indicators that are used to calculate the IMD score. These domains and indicators contain a richer set of data that might be useful for classifying local authorities. Clustering methods offer a set of techniques to identify groups of areas with similar patterns of deprivation. This could offer insights into areas that face similar challenges.

**Methods:** Hierarchical agglomerative (i.e. bottom-up) clustering methods were applied to sub-domain scores for 152 upper-tier local authorities. Recent advances in statistical testing allow clusters to be identified that are unlikely to have arisen from random partitioning of a homogeneous group. The resulting clusters are described in terms of their subdomain scores and basic geographic and demographic characteristics.

**Results:** Five statistically significant clusters of local authorities were identified. These clusters represented local authorities that were:

i. Most deprived, predominantly urban;
ii. Least deprived, predominantly rural;
iii. Less deprived, rural;
iv. Deprived, high crime, high barriers to housing; and
v. Deprived, low education, poor employment, poor health.

**Conclusion:** Hierarchical clustering methods identify five distinct clusters that do not correspond closely to quintiles of deprivation. These methods can be used to draw on the richer set of information contained in the IMD domains and may help to identify places that face similar challenges, and places that appear similar in terms of IMD scores, but that face different challenges.

## INTRODUCTION

The English Indices of Multiple Deprivation (IMD) is a commonly used measure of relative deprivation for geographic areas in England. It is often used for analysing inequalities in health between more and less deprived areas.

The IMD provides an overall score of relative deprivation that is used to rank Lower-Super Output Areas (LSOAs, small geographic areas in England, each with a population of approximately 1,500). The overall score is calculated as a weighted sum of seven domains of deprivation. These domains measure deprivation in: income; employment; health and disability; education, training and skills; crime; access to housing and services; and living environment. Each of the domains is made from one or more indicators. Where more than one indicator is used, indicators are combined using weights determined by factor analysis to create the subdomain score. The weights used to combine the domains into the overall score were determined using a range of methods including previous research that the income and employment domains should be weighted more heavily, as well as empirical research on public preferences and preferences revealed by relative levels of government spending on each domain (Smith et al. 2015).

Summary IMD scores are available for larger geographic areas, such as upper-tier local authorities (the upper-most layer of local government in England). For each upper-tier local authority, a number of different ways of combining the deprivation of its LSOAs are used, including average IMD score, average rank, and the proportion of population living in the 10% most deprived LSOAs (Smith et al. 2015).

In analyses at local authority level, each local authority is typically ranked according to the average score or average rank of its constituent LSOAs, and local authorities are then grouped into quintiles or deciles. This then forms the basis for estimating inequalities in health outcomes. For example, the Public Health Outcomes Framework, a widely used tool for understanding variations in health between English local authorities, has an option to view health indicators by decile of deprivation (Public Health England 2018).

While not wholly arbitrary, the method of grouping areas together in quintiles or deciles may group together areas with quite different deprivation profiles. For example, when upper-tier local authorities are ranked by their average IMD scores, North East Lincolnshire and South Tyneside are ranked as the 25th and 26th most deprived local authorities respectively (out of 152 upper-tier local authorities), both in the second most deprived decile. However, their scores on the domains are different: in terms of average health and disability deprivation South Tyneside is ranked 12th most deprived (in the most deprived decile), while North East Lincolnshire is ranked 66th most deprived (in the fifth most deprived decile). As a result, the use of a single overarching ranking system may obscure differences in the types of challenges faced by local authorities, and does not make full use of the rich information contained in the full set of sub-domain scores.

Statistical clustering methods offer a way to find groups of geographic areas that face similar challenges. For example, Bellis et al (2012) used k-means clustering on health outcomes data and found that the cluster with the worst health outcomes was concentrated in the ex-industrial areas of the North of England.

An alternative to the k-means approach is hierarchical clustering, which produces a set of nested clusters, allowing the researcher to describe the clustering structure in the data without arbitrarily specifying a number of clusters to be found. Recent advances in statistical methods make it possible to assess at each ‘branch’ in the tree, whether the resulting clusters are likely to have arisen by chance or not, allowing a clustering structure to be determined without the researcher pre-determining how many clusters will be found (Kimes et al. 2017).

This complements the use of ‘nearest neighbours’ models, such as that produced by the Chartered Institute of Public Finance and Accounting (CIPFA 2017). This model calculates the Euclidean distance between a given local authority and all other local authorities based on a selection of indicators. This approach is designed to find similar local authorities given an initial local authority, rather than grouping all local authorities based on patterns of deprivation.

The aim of this paper is to apply clustering techniques to domains of deprivation to explore patterns of deprivation among upper-tier local authorities in England. It aims to test the usefulness of unsupervised statistical learning methods in understanding the challenges faced by local authorities and to find out if any resulting clusters are consistent with a single continuum of multiple deprivation, or whether there are clusters that differ more in the pattern of deprivation than in their overall IMD scores.

## METHODS

### Data sources and processing

IMD data were downloaded from the website of the Ministry of Housing, Communities and Local Government (Ministry of Housing, Communities and Local Government 2015). Geographic data were downloaded from the Office for National Statistics Open Geography Portal (geoportal.statistics.gov.uk/datasets/). Urban rural classification data were downloaded from the Office for National Statistics. Demographic data was downloaded using the FingertipsR package (Fox et al. 2017).

The IMD summary statistics for upper-tier local authorities include a range of scores that summarise the IMD scores of each local authority’s component LSOAs. These include the average of LSOA scores in each sub-domain, and the proportion of LSOAs in a given local authority in the lowest 10% nationally. For simplicity, this study uses only the average scores for each local authority for the seven domains of deprivation. Average scores for each local authority were centered and scaled to mean 0 and standard deviation of 1. This prevents the distribution of any variable affecting its weight in the clustering analysis.

### Statistical analysis

This study uses a hierarchical agglomerative (i.e. bottom-up) clustering algorithm. The algorithm computes the Euclidean distance between each local authority based on average scores on the seven domains of deprivation. The algorithm then identifies the two local authorities that have the lowest distance score and links them in a cluster. The algorithm then repeats this process until only one cluster remains containing all the local authorities. The ‘complete’ linkage method was used, where the distance between two clusters is the distance between their two farthest points.

Statistical analysis was done in R version 3.5.0 (R Development Core Team 2008). Hierarchical clustering was done using the sigclust2 package (Kimes et al. 2017). This package enables testing for statistical significance of clustering. At each node, the algorithm computes the two-mean cluster index, a measure of the extent to which observations within the two clusters vary relative to the overall variation in the data set (small values indicating tighter clustering). The value of the cluster index at each node is then compared to the distribution expected under the null hypothesis that both clusters are drawn from a single gaussian distribution. This produces a p-value for each node in the clustering structure indicating the likelihood that a cluster index that large or larger could have arisen by chance. To be conservative, nodes with p-values less than 0.001 were treated as statistically significant. To correct for multiple testing, the algorithm applies a family-wise error rate correction, which shrinks the critical value at which the clustering at a given node is judged to be statistically significant. If a node has a p-value greater than 0.001, the algorithm does not test nodes below it in the clustering hierarchy. Using this approach, the number of clusters identified is determined by the degree of clustering, rather than being decided in advance by the researcher, or being based on an arbitrary cut-off point on the clustering tree.

As the sub-domain scores were used to create the clustering, testing for statistically significant differences between cluster subdomain scores is neither necessary nor appropriate. Several local authority domains and other variables were skewed. For consistency medians and interquartile ranges are reported, and non-parametric statistical tests are used. Cluster geographic and demographic characteristics were compared using Kruskal-Wallis tests. Where the Kruskal-Wallis test for a given cluster characteristic was significant at p < 0.05 post-hoc Dunn’s tests using Bonferroni correction for multiple comparisons were used to identify statistically significant differences between pairs of clusters.

All R code is available online (see supplementary methods).

## RESULTS

### Hierarchical clustering

Figure 1a shows the clustering structure as a dendrogram. Statistically significant clustering is identified by red branches in the dendrogram, and associated p-values are displayed where the clustering at that node was statistically significant. Five statistically significant clusters were identified. Figure 1b shows the geographic distribution of the five significant clusters.

**Figure 1:**
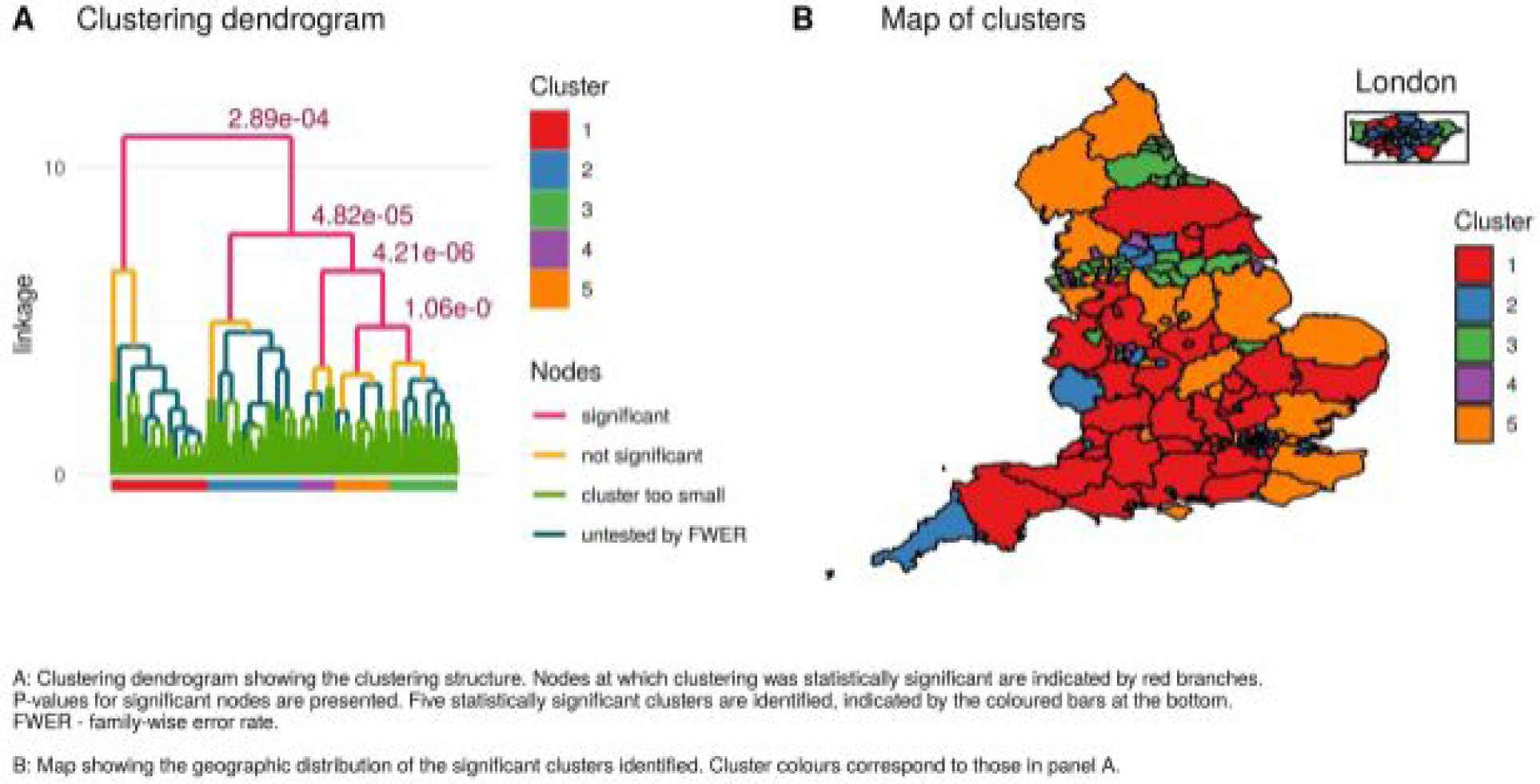
Heirarchical clustering of local authorities by subdomains of deprivation.

Figure 2 and table 1 show the median subdomain z-scores scores for each of the clusters. Table 2 shows how the local authorities are distributed across the clusters and quintiles of IMD scores.

**Figure 2:**
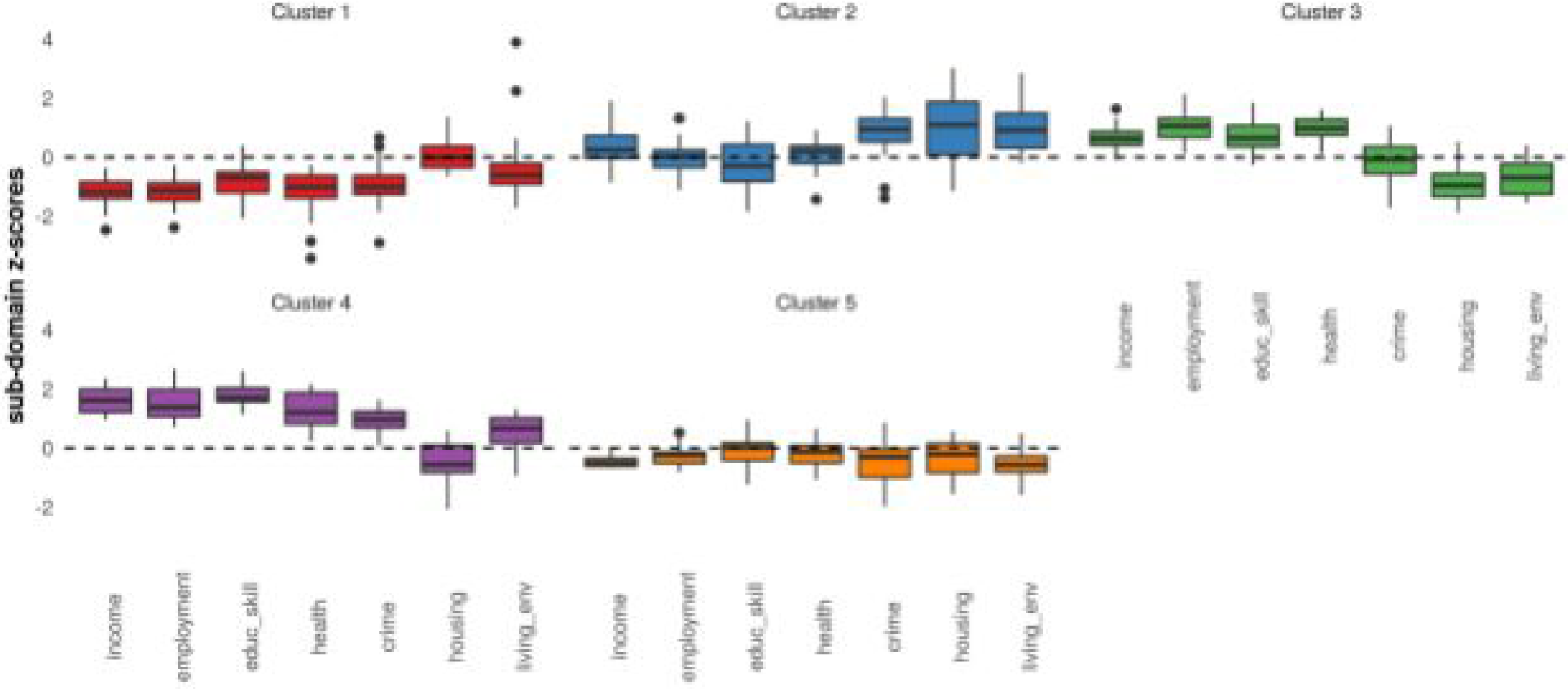
Cluster deprivation profiles. Median IMD subdomain scores by cluster

**Table 1:** cluster subdomain scores.

| Table 1: cluster subdomain scores |  |  |  |  |  |  |  |  |  |
| --- | --- | --- | --- | --- | --- | --- | --- | --- | --- |
| Cluster | n | Median<br>IMD<br>score | Subdomain z-scores: median (interquartile range) |  |  |  |  |  |  |
|  |  |  | Income | Health | Crime | Employ<br>ment | Educat<br>ion and<br>skills | Housin<br>g | Living<br>environ<br>ment |
| 1 | 42 | 13.81<br>(3.59) | -1.17<br>(0.56) | -1.12<br>(0.6) | -0.69<br>(0.7) | -1 (0.76) | -1<br>(0.63) | 0 (0.73) | -0.6 (0.7) |
| 2 | 41 | 26.64<br>(5.01) | 0.24<br>(0.78) | 0.03<br>(0.59) | -0.28<br>(1.24) | 0.21<br>(0.58) | 0.95<br>(0.79) | 1.1 (1.8) | 0.92<br>(1.18) |
| 3 | 30 | 28.27<br>(3.95) | 0.63<br>(0.46) | 1.07<br>(0.66) | 0.65<br>(0.72) | 0.98<br>(0.53) | -0.06<br>(0.93) | -0.94<br>(0.79) | -0.69<br>(1.04) |
| 4 | 15 | 34.61<br>(7.64) | 1.62<br>(0.77) | 1.38<br>(0.92) | 1.71<br>(0.46) | 1.21<br>(1.07) | 0.97<br>(0.56) | -0.55<br>(0.93) | 0.66<br>(0.85) |
| 5 | 24 | 18.84<br>(2.76) | -0.5<br>(0.28) | -0.23<br>(0.37) | 0.02<br>(0.57) | -0.14<br>(0.58) | -0.31<br>(0.95) | -0.21<br>(0.98) | -0.57<br>(0.53) |

**Table 2:**
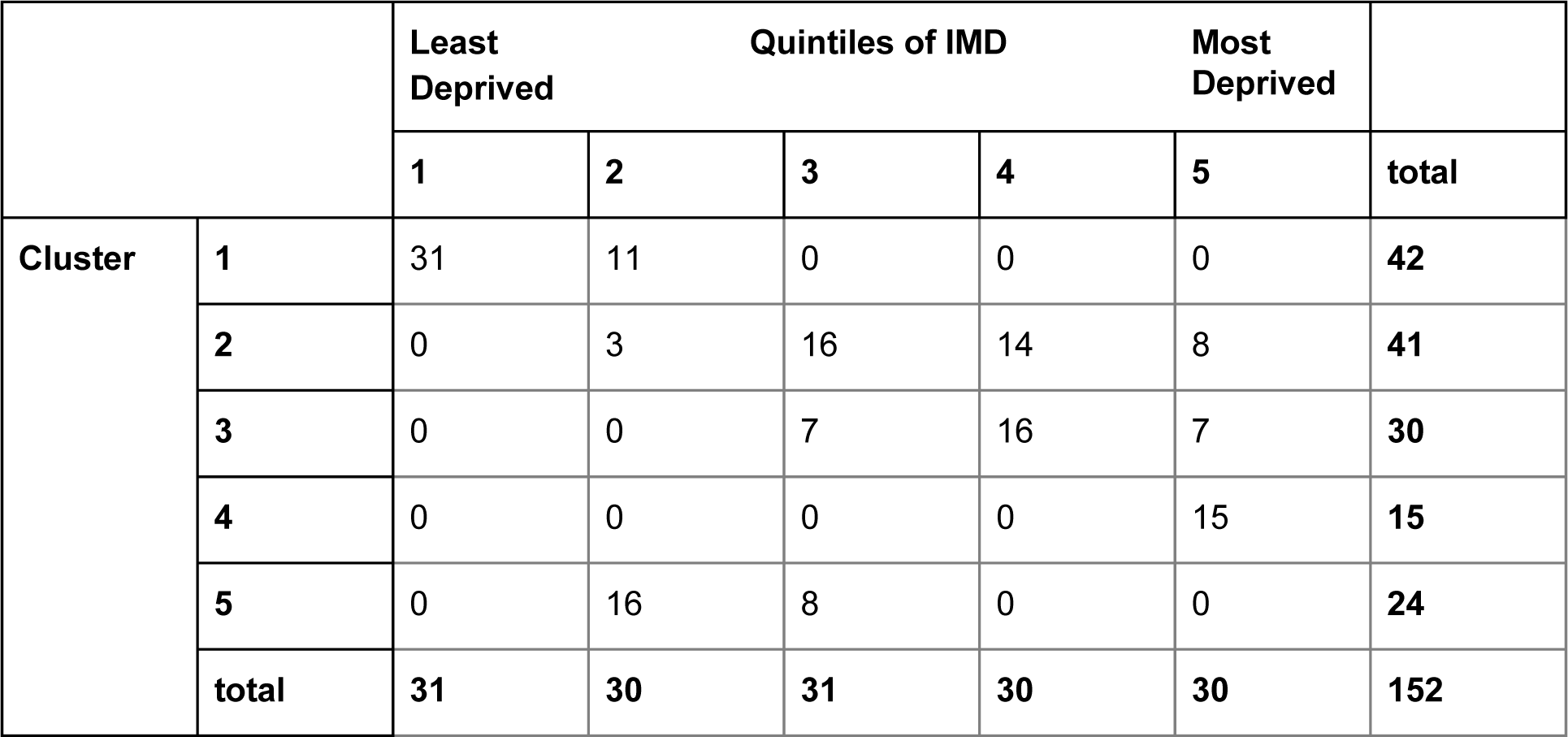
distribution of clusters across quintiles of IMD.

The clusters form a nested structure described by the dendrogram shown in figure 1 panel A. Clusters 2, 3, 4, and 5 are nested within a larger cluster that is distinct from cluster 1. This suggests that there is more difference in terms of patterns of deprivation between cluster 1 (the least deprived cluster) and the remaining clusters. Looking further down the dendrogram, cluster 2 is separated from clusters 3, 4, and 5. The profiles of IMD subdomain scores in figure 2 suggest that cluster two has a different pattern of deprivation to clusters 3, 4, and 5. Cluster 4 is then separated from clusters 3 and 5, which are the last to be separated.

Cluster 1, the least deprived overall, contains 42 local authorities. Average scores for local authorities in this cluster are low on all domains of deprivation apart from barriers to housing, which are around average. These areas are mainly large rural counties, with some more affluent boroughs of London.

Cluster 2, comprising 41 local authorities, has average deprivation scores only slightly above average for income, health, and employment, and slightly below average deprivation in education, but high levels of deprivation in crime, housing, and living environment. These areas include a mix of London boroughs as well as some more rural areas, such as Kirklees and Cornwall.

Cluster 3 (30 local authorities) has a similar average IMD score to cluster 2 but experiences low levels of housing and environmental deprivation and average crime levels, but high deprivation in income, employment, health, and education and skills. These areas are mainly post-industrial towns concentrated in the North of England.

Cluster 4 is the most deprived overall. Average deprivation scores are high across all domains except housing. This cluster is relatively small, containing only 15 local authorities, mostly deprived towns and cities in the North and Midlands, such as Blackpool, Liverpool, Manchester, and Wolverhampton.

Cluster 5 (24 local authorities), the second least deprived, scores slightly below average across all domains of deprivation, with lower scores in income and environmental deprivation. This cluster includes a mix of rural and urban local authorities, many county councils, such as Northamptonshire and Lincolnshire, which contain both relatively deprived and relatively affluent areas.

Table 2 shows the distribution of the clusters across quintiles of deprivation. There are extensive overlaps between the clusters’ distributions across the quintiles of deprivation. This suggests that the clusters do not correspond to a simple continuum of multiple deprivation. Only cluster 4 is entirely contained within a single quintile - in this case the most deprived quintile. However cluster 4 only comprises half of the most deprived local authorities. The remainder fall in clusters 2 and 3. Local authorities in cluster 1 fall entirely in the least two deprived quintiles. There is extensive overlap between clusters 2 and 3, with most local authorities in each cluster falling in IMD quintiles 3, 4, and 5 (i.e. the three most deprived quintiles).

Figure 3 shows how the clusters of local authorities differ in geographic size, age structure and the proportion of the population that lives in areas classified as rural by the ONS. Table 3 reports the results of Kruskall-Wallis tests for statistically significant differences between clusters.

**Figure 3:**
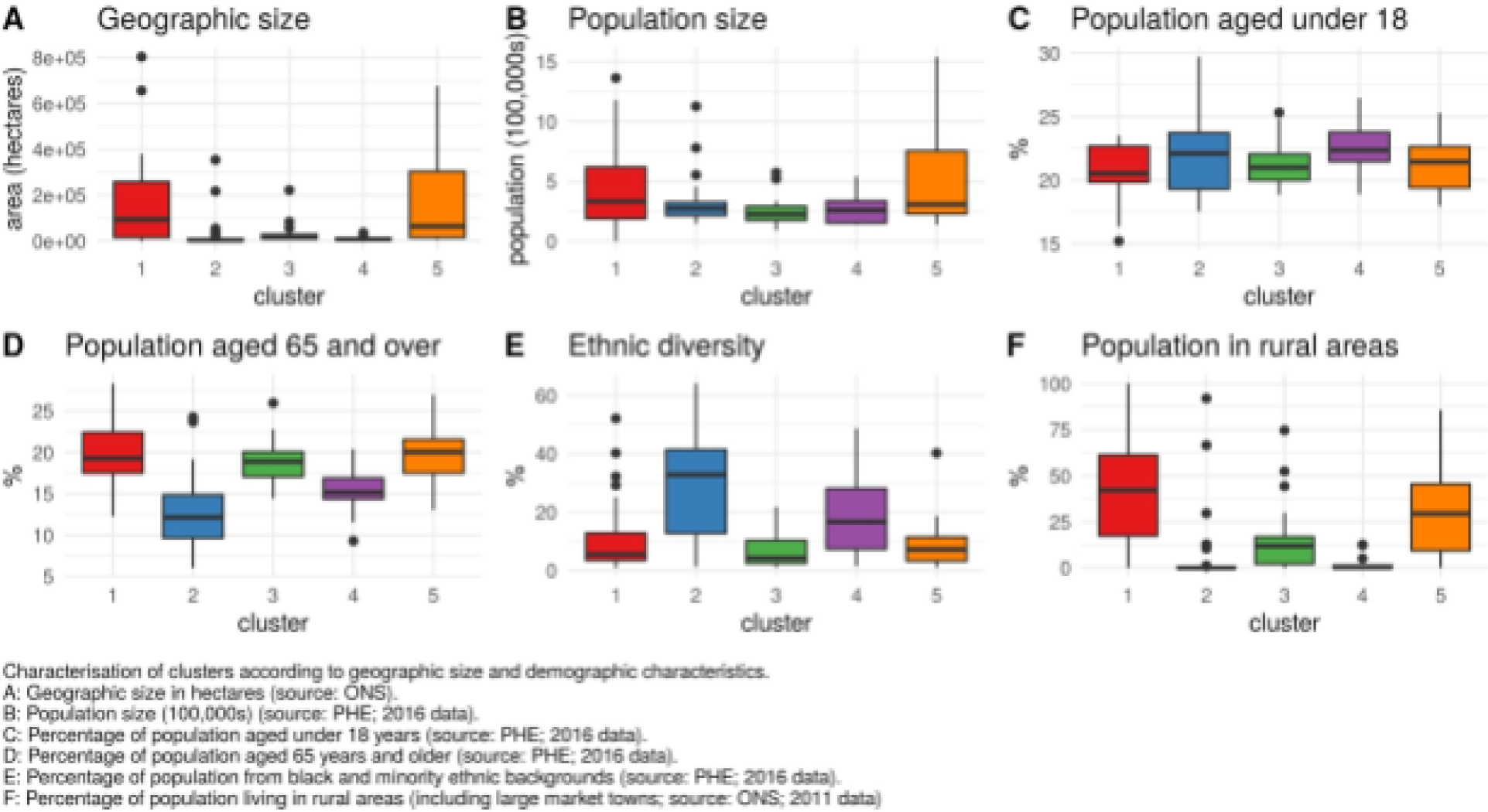
Cluster characteristics.

**Table 3:** Cluster characteristics.

| Variable | Cluster median (interquartile range) |  |  |  |  | H | df | p-value |
| --- | --- | --- | --- | --- | --- | --- | --- | --- |
|  | 1 | 2 | 3 | 4 | 5 |  |  |  |
| Area (10,000 HA) | 9.41<br>(24.32) | 0.43<br>(0.5) | 1.55<br>(1.84) | 0.86<br>(0.41) | 6.48<br>(28.89) | 55.59 | 4 | < 0.001 |
| Total population (100,000s) | 3.33<br>(4.31) | 2.74<br>(1.11) | 2.28<br>(1.16) | 2.6<br>(1.83) | 3.09<br>(5.25) | 11.37 | 4 | 0.023 |
| % aged under 18 | 20.54<br>(2.82) | 22.12<br>(4.42) | 21.01<br>(2.07) | 22.37<br>(2.31) | 21.44<br>(3.24) | 9.18 | 4 | 0.057 |
| % aged 65 and over | 19.27<br>(4.93) | 12.12<br>(5.21) | 18.87<br>(2.99) | 15.15<br>(2.53) | 20.03<br>(4.1) | 61 | 4 | < 0.001 |
| % ethnic minority | 5.5 (9.4) | 32.8<br>(28.7) | 4.25<br>(7.85) | 16.6<br>(20.75) | 7.15<br>(8.25) | 46.2 | 4 | < 0.001 |
| % living in rural areas | 42.13<br>(43.92) | 0 (0.17) | 11.87<br>(14.68) | 0.21<br>(1.23) | 29.41<br>(36.25) | 68.2 | 4 | < 0.001 |

Clusters 1 and 5 contain most of the English counties, and as such tend to be significantly larger than clusters 2 and 4 which contain mostly unitary authorities (Kruskall-Wallis statistic 55.59 on 4 degrees of freedom, p < 0.001; post-hoc Dunn’s tests for pairwise comparisons between cluster 1 and clusters 2 and 4, and between cluster 5 and clusters 2 and 4 all significant at p < 0.05).

The same pattern is seen in the proportion of the population who live in rural areas, with clusters 1 and 5 having significantly more of their population in areas classified as rural than clusters 2 and 4 (Kruskall-Wallis statistic 68.2 on 4 degrees of freedom, p < 0.001, Dunn’s tests for pairwise comparisons between clusters 1 and clusters 2 and 4 and cluster 5 and clusters 2 and 4 significant at p < 0.001).

There is some evidence that clusters vary in size of population. (Kruskall-Wallis statistic 11.37 on 4 degrees of freedom, p = 0.023). However in pairwise comparisons, only clusters 3 and 5 differed significantly (Dunn’s tests for pairwise comparisons significant at p = 0.025).

There was limited evidence of differences in the proportion of the population aged under 18 (Kruskall-Wallis test statistic 9.18 on 4 degrees of freedom, p = 0.057), but clusters did appear to differ in the proportion of the population aged 65 and over (Kruskall-Wallis test statistic 61 on 4 degrees of freedom, p < 0.001). Cluster 2 had the lowest proportion aged 65 plus (Dunn’s test comparisons between cluster 2 and clusters 1, 3, and 5 all significant at p < 0.05), followed by cluster 4, which differed significantly from clusters 1 and 5 (post-hoc Dunn’s tests significant at p < 0.001).

Clusters also varied in ethnic diversity (Kruskall-Wallis test statistic 46.2 on 4 degrees of freedom, p < 0.001). Cluster 2 was the most ethnically diverse, with clusters 1, 3, and 5 less ethnically diverse (Dunn’s tests for pairwise comparisons between clusters 2 and 1, 3, and 5 significant at p < 0.001). Cluster 4 had a higher median ethnic diversity, but pairwise comparisons with other clusters were not statistically significant at the p < 0.05 level.

## DISCUSSION

### Key findings

Hierarchical clustering methods identify five statistically significant clusters of local authorities. These clusters are different groupings from the quintiles of deprivation commonly used. This suggests that hierarchical clustering methods offer an alternative way to explore patterns of deprivation at local authority level. The identification of clusters of local authorities that share patterns of deprivation may help to guide public health and other policy interventions. For example, successful interventions in one local authority might transfer more easily to other areas within the same cluster, as these areas may share similar challenges and other contextual features.

While three of the clusters reported here appear to reflect groups of local authorities with low, medium, and high levels of deprivation across all IMD domains, clusters 2 and 3 have similar overall deprivation scores, but have contrasting patterns of deprivation. This suggests that the use of the overall IMD score to group local authority areas obscures some of the information contained in the IMD subdomain scores, and groups together areas that face different challenges.

Local authorities in cluster 2 tend to have higher than average deprivation scores for crime, housing, and living environments, but average deprivation scores for the other IMD domains. Cluster 3 has the opposite pattern: higher than average scores on income, employment, education and health, but average scores on crime, housing, and living environment deprivation. Importantly for public health policy and practice, local authorities in cluster 3 appear to have worse health deprivation than their overall deprivation scores might predict. These differences may be explained by some of the differences in the characteristics of these clusters of local authorities shown in figure 3. Cluster two appears to contain local authorities whose populations are on average more educated, less likely to be retired, and more ethnically diverse, and more urban than those in cluster 3. This suggests that it is important to look at the pattern of deprivation and the social and demographic factors underlying it, not only the overall IMD score.

The fact that local authorities in cluster 3 are concentrated in Northern England may help to explain the observation that areas in the North of England appear to suffer worse health even after correcting for IMD score (Whitehead et al. 2014; Kontopantelis et al. 2018). This suggests that the construction of the IMD may not be accurately capturing deprivation that affects health, and may under-weight deprivation in Northern areas of England.

The results presented here suggest that the use of quintiles or deciles of deprivation for benchmarking local authority performance against may be misleading. The statistical neighbours approach (for example that used by CIPFA) may be better able to identify comparable local authority areas (CIPFA 2017).

### Strengths and limitations

#### Strengths

The combination of hierarchical clustering methods and an approach to identifying statistically significant clustering provides a less arbitrary approach to grouping local authorities. The five statistically significant clusters identified here display a degree of face-validity (reflecting areas with similar economic histories), while adding value by identifying similarities and groupings that are not captured when local authorities are grouped based on their overall IMD score. This offers a more nuanced picture of patterns of deprivation among local authorities in England.

#### Limitations

The use of upper-tier local authorities in this analysis is intended to make the results useful for local authority and national policymakers. However, it means that the resulting clustering may be affected by the large variation in types of local authority. The use of English upper-tier local authorities also limits the sample size to 152. Further analysis using smaller areas such as LSOAs would partly remove variation in population size, and would substantially increase the statistical power of the analysis, potentially allowing for finer distinctions between patterns of deprivation. Such an analysis might also help to reveal similarities between small areas in different local authorities. This could help to spread successful local interventions by finding other areas that share similar challenges and contexts.

Another further extension to this work would be to test the stability of the clustering structure over time. The recent publication of the 2019 Indices of Deprivation (Noble et al. 2019) along with the earlier 2010 Indices of Deprivation (DCLG 2011) provide a longitudinal data set with a largely unchanged underlying set of indicators. Analysing the clustering structure over time would test whether the patterns of deprivation described here represent enduring features of English social geography.

It should also be noted that the results may be sensitive to decisions about which variables to use. The decision to use only average scores for the seven domains is also likely to have influenced the resulting clustering structure. The results are also sensitive to the choice of distance and linkage methods. Further work is needed to provide a sound basis for these analytical choices.

## Data Availability

The manuscript is based entirely on publicly available data.

https://github.com/stevenlsenior/IMD_clustering

## Notes

### Competing Interest Statement

The authors have declared no competing interest.

### Funding Statement

No funding was received for this work.

### Author Declarations

All relevant ethical guidelines have been followed and any necessary IRB and/or ethics committee approvals have been obtained.

Any clinical trials involved have been registered with an ICMJE-approved registry such as ClinicalTrials.gov and the trial ID is included in the manuscript.

